# Increased mortality associated with uncontrolled diabetes mellitus in patients with Pulmonary Cryptococcosis — a single U.S. cohort study

**DOI:** 10.1101/2021.01.11.21249631

**Authors:** Solana Archuleta, Amal A. Gharamti, Stefan Sillau, Paula Castellanos, Sindhu Chadalawada, William Mundo, Mehdi Bandali, Jose Oñate, Ernesto Martínez, Daniel Chastain, Kristen DeSanto, Leland Shapiro, Ilan S. Schwartz, Carlos Franco-Paredes, Andrés F. Henao-Martínez

## Abstract

**Background:** Diabetes mellitus is an established risk factor for bacterial infections, but its role in Cryptococcosis is unclear. The study aimed to determine whether uncontrolled diabetes (HbA1c >7%) was an independent risk factor for mortality in cryptococcosis.

**Methods:** A retrospective case-control study partially matched by age and gender was performed in patients tested for *Cryptococcus* infection at the University of Colorado Hospital from 2000-2019. A multivariable logistic regression model was used to identify mortality predictors. Cox proportional hazard model was used for survival analysis.

**Results:** We identified 96 cases of Cryptococcosis and 125 controls. Among cases, cryptococcal meningitis (49.0%) and pneumonia (36.5%) constituted most infections. Cases with pulmonary cryptococcosis had a higher mortality at 10 weeks (50% vs 7%, p=0.006) and one year (66.7% vs 13.8%, p=0.005). Unadjusted Cox proportional hazard model found an increased rate of death for uncontrolled diabetes at 10-weeks (hazard ratio 8.4, CI: 1.4-50.8, p=0.02), and 1-year (hazard ratio 7.0, CI: 1.7-28.4, p=0.007) among pulmonary cryptococcosis cases. Multivariable analysis showed a significantly increased odds of 10-weeks (OR=4.3, CI: 1.1-16.5, p=0.035) and one-year (OR=5.9, CI: 2.2-15.8, p=0.014) mortality for uncontrolled diabetes among pulmonary cryptococcosis cases. After adjustment for gender, age, and case/control, for every 1% increase in HbA1c levels, the odds of pulmonary cryptococcosis mortality at one-year increased by 11% (OR = 1.6, CI 95%: 1.1-2.3, p= 0.006).

**Conclusion:** Uncontrolled diabetes is associated with worse outcomes in pulmonary cryptococcosis, including a 4-fold and 6-fold increased odds of death at 10-weeks and 1-year, respectively. Glucose control interventions should be explored to improve clinical outcomes in patients with pulmonary cryptococcosis.

## Introduction

Diabetes mellitus (diabetes) is one of the most prevalent non-communicable diseases in the United States. The American Diabetes Association estimates 34.2 million Americans (10.5% of the population) are affected (1), and the prevalence increases with age. Among those aged 65 or older, diabetes is present in 26.8%. A common complication among patients with diabetes is infections, which carries a higher risk of mortality compared to adults without diabetes (2). Pneumonia and sepsis are the most common infections associated with mortality in diabetes. The pathogenesis of hyperglycemia-induced immune dysfunction includes reduced neutrophil chemotaxis, adhesion, and migration, and decreased intracellular production of hydrogen peroxide (3, 4). Diabetes is also a known risk factor for invasive fungal infections, such as mucormycosis (5). However, we lack reports of diabetes or increased glucose levels as independent risk factors for opportunistic infections commonly affecting advanced immunocompromised states, including *Cryptococcus*.

Cryptococcosis continues to be a source of morbidity and mortality —up to 20% — among immunocompromised hosts (6). Cryptococcal meningitis has a significant global impact with 181,100 deaths each year among HIV-infected individuals (7). Moreover, developing countries have seen an increasing incidence of the disease among non-HIV immunocompromised hosts as well (8, 9). Solid-organ transplantation, systemic lupus erythematosus, malignancy, sarcoidosis, and cirrhosis are conditions of functional immunosuppression known to increase the risk for *Cryptococcus* spp. infection (10-15). Diabetes mellitus is often listed as an additional risk factor for Pulmonary cryptococcosis especially among HIV-negative patients (16, 17).

Diabetes is a common comorbidity among adult patients in the U.S. with Cryptococcosis. However, the role of diabetes in the prevalence and associated mortality of *Cryptococcus* infections is not well defined. The study aimed to determine whether diabetes and HbA1c levels were independent risk factors for mortality in *Cryptococcus* infection.

## Methods

### Ethics statement

The present investigation is in the Health Insurance Portability and Accountability Act (HIPAA) compliance and was approved by the Colorado Multiple Institutional Review Board (COMIRB) at the University of Colorado Denver. Analysis of clinical data has been performed under an approved protocol (COMIRB Protocol 15-1340), and an exemption of informed consent was granted.

### Patients and data collection

We collected data on all patients with culture from any site that grew *Cryptococcus* or with detection of antigens from serum or cerebrospinal fluid [CSF] diagnosed at the University of Colorado Hospital microbiology laboratory between January 2000 and October 2019.

Controls were randomly chosen among patients with negative serum and when done, CSF cryptococcal antigen and culture. A manual chart review was performed to exclude clinical diagnoses of Cryptococcosis in these patients. Controls were partially matched to cases by specimen collection date, age, and biological sex. Controls were mainly hospitalized and also ambulatory patients from the same community served by the University of Colorado Hospital. Most control patients had suspicions of having Cryptococcosis based on their underlying immunosuppressive state and clinical concerns for meningitis or abnormal pulmonary findings.

The cryptococcal antigen tests used were an enzyme immunoassay test (Meridian Premier Cryptococcal assay) before 2013, and the lateral flow assay (CrAg® LFA, Immuno-Mycologics Inc. [IMMY], Norman, OK) after 2013. Cryptococcal meningitis was diagnosed based on a positive cryptococcal CSF antigen or positive CSF culture. The assay didn’t differentiate the species of *Cryptococcus*. Medical reports were manually accessed to collect clinical and laboratory variables for all patients. The following data was retrospectively collected through RedCap (electronic data capture tools hosted at the University of Colorado Denver): demographics (gender, race, and age); symptoms (constitutional, headaches, altered mental status, respiratory abnormalities, fever, and others), medical history (smoking, lung disease, diabetes, hypertension, lupus, malignancy, sarcoidosis, cirrhosis, HIV infection, solid organ transplant, use of calcineurin inhibitors or corticosteroids, and prednisone dose); transplant (type and time since transplant); vital signs at collection times (systolic blood pressure, diastolic blood pressure, pulse, and temperature); weight, and body mass index (BMI); diagnosis of cryptococcal meningitis; laboratory results (complete blood cell count, comprehensive metabolic panel, baseline renal function, and blood culture), and outcomes of cryptococcal infection: 10-weeks, 1-year, and overall mortality (per consensus criteria established by the Mycoses Study Group and European Organization for Research and Treatment of Cancer (18)), cognitive deficits, muscle weakness, speech difficulties, hearing impairment, stroke by MRI results; and use of ventriculoperitoneal shunts (VPS). Uncontrolled diabetes was defined as the most recent HbA1c level>7%. Electronic medical records (EPIC, Verona, WI) were automatically interrogated for A1C levels and date of death from the cohort of patients through software supported by Health Data Compass Data Warehouse project (healthdatacompass.org). The most recent HbA1c available before admission was chosen for analysis. The vast majority of patients with a diagnosis of diabetes had an HbA1c level available.

### Statistical analysis

Statistical analyses were performed using STATA software, version 12.1 (StataCorp, College Station, Texas, USA). We had crude mortality as our primary outcome at 70 days, and one year. The means and standard deviations for continuous variables were calculated. For categorical variables, frequencies and percentages were calculated. We initially performed a bivariate analysis for dichotomous outcomes variables using chi-squared, and the Fisher exact test for dichotomous and nominal independent variables, respectively. For interval independent variables, we used the t-test. The uncontrolled diabetes group was compared to patients with controlled diabetes (HbA1c≤7%) or no diabetes. For the multivariable analysis, we selected age, sex, commonly known cryptococcosis risk factors (HIV, solid organ transplant, cirrhosis, malignancy, and steroids), smoking, factors associated with increased mortality in Cryptococcosis (altered mental status and positive blood cultures for Cryptococcus), and case vs control status. We did not include co-linear variables. Selected variables were included in a multivariable, forward, stepwise logistic regression model. A parallel conditional logistic regression model was run for comparison. Kaplan–Meier survival curves were constructed to show cumulative mortality over the study period for Cryptococcosis cases by uncontrolled diabetes mellitus status. Cox regression was used to estimate the mortality rate ratio among pulmonary cryptococcosis cases with and without uncontrolled diabetes. Due to heterogeneous controls, we also run a propensity score matching analysis. Linearity between HbA1c and days from diagnosis to death was checked using a scatter plot and R-squared/adjusted R-squared from ANOVA.

### Data access

The corresponding author had full access to data in the study and had final responsibility for the decision to submit the manuscript for publication. The datasets generated and analyzed in the current study are available from the corresponding author on reasonable request.

## Results

We included a total of 96 cases and 125 controls. At least 55 cases and 112 controls were matched by specimen collection date, age, and gender.

### Clinical characteristics of patients with Cryptococcosis

Most subjects were men (79%). The average age was 54.1 years, and most subjects were Caucasian with an upper limit of normal BMI. Diabetes was present in about a quarter of the cases, with no statistical difference with controls. Among cases, cryptococcal meningitis (49.0%) and pneumonia (36.5%) constituted most infections; followed by skin (8.3%), and asymptomatic antigenemia (6.3%). Other common risk factors among cases were HIV (38.9%), steroid use (24.7%), malignancy (21.1%), solid organ transplantation (18.1%), and cirrhosis (5.2%). Overall mortality was similar among cases and controls (38.5% vs. 39.2%, p=0.921).

### Clinical characteristics of patients with Pulmonary Cryptococcosis

Pulmonary cryptococcosis cases (n=35) were older and were more likely to be HIV negative (table 1). They commonly presented with respiratory symptoms as opposed to headaches and altered mental status. Controls shared similar demographics and risk factors. Mean hemoglobin and platelet levels were within or close to normal limits and similar among the two groups. Creatinine levels were similar among cases and controls. Although HbA1c levels were higher among pulmonary cryptococcosis cases, they were not statistically significant. ICU stay, overall mortality, and mortality at 10 weeks and one year were similar to controls.

**Table 1.** Patient clinical characteristics for cases and controls.

| Patient characteristics<br>(%, mean ± SD) | N | Controls (n=125) | Pulmonary cryptococcosis<br>cases (n=35) | P-values |
| --- | --- | --- | --- | --- |
| **Demographics** |  |  |  |  |
| Gender (male) | 221 | 106 (84.8%) | 27 (77.1%) | 0.285 |
| Age (years) | 221 | 53.6 ± 14.9 | 59.5 ± 14.5 | 0.038 |
| Race (white) | 126 | 73 (63.5%) | 22 (64.7%) | 0.242 |
| BMI (kg/m<sup>2</sup>) | 147 | 25.3 ± 5.9 | 24.9 ± 4.7 | 0.757 |
| **Risk factors** |  |  |  |  |
| Smoking (current) | 111 | 30 (24.4%) | 4 (11.4%) | 0.223 |
| Transplant | 219 | 28 (22.4%) | 9 (26.5%) | 0.618 |
| Diabetes mellitus | 221 | 25 (20%) | 9 (25.7%) | 0.465 |
| Uncontrolled DM | 211 | 9 (7.6%) | 6 (17.1%) | 0.093 |
| HIV | 220 | 46 (36.8%) | 6 (17.7%) | 0.035 |
| Malignancy | 220 | 31 (24.8%) | 12 (35.3%) | 0.222 |
| Cirrhosis | 221 | 6 (4.8%) | 1 (2.9%) | 0.619 |
| Steroids | 217 | 27 (21.8%) | 9 (27.3%) | 0.504 |
| **Symptoms at presentation** |  |  |  |  |
| Headaches | 219 | 33 (26.6%) | 2 (5.9%) | 0.010 |
| Altered mental status | 221 | 48 (38.4%) | 4 (11.4%) | 0.003 |
| Respiratory | 220 | 32 (25.8%) | 28 (80%) | < 0.001 |
| **Labs** |  |  |  |  |
| Hemoglobin (g/dL) | 215 | 12.3 ± 2.6 | 11.9 ± 2.9 | 0.513 |
| Platelets (10<sup>9</sup>/L) | 214 | 207.9 ± 135.2 | 207.7 ± 84 | 0.992 |
| Creatinine (mg/dl) | 208 | 1.3 ± 1.2 | 1.3 ± 0.8 | 0.843 |
| CD4 count (cells/μL) | 86 | 206 ± 224.9 | 200.9 ± 319.1 | 0.952 |
| HbA1c (%) | 96 | 6.24 ± 1.8 | 6.8 ± 1.7 | 0.252 |
| **Outcome** |  |  |  |  |
| ICU stay | 53 | 27 (36%) | 7 (30.4%) | 0.624 |
| Overall Death | 221 | 49 (39.2%) | 15 (42.9%) | 0.696 |
| Death within 10 weeks | 221 | 12 (9.6%) | 5 (14.3%) | 0.427 |
| Death within 1 year | 221 | 28 (22.4%) | 8 (22.9%) | 0.954 |

### Clinical factors associated with an increase of 10-weeks and 1-year mortality from Cryptococcosis

We found 10-weeks and 1-year mortality of 19% and 22% among cryptococcosis cases. The 10-weeks mortality was significantly higher in cases compared to controls (18.8% vs 9.6%, p=0.049). At 10 weeks, diabetes mellitus (44.4% vs 21.8%, p=0.048) and uncontrolled diabetes (31.3% vs 7.9%, p=0.009) were more commonly present in those who died. Conversely, transplant recipients are less likely to die shortly after diagnosis (5.6% vs 21%, p=0.001). Altered mental status, anemia, and lower mean platelet count associate with an increase in 10-weeks mortality (table 2). At 1-year, transplant recipients maintained lower mortality compared to non-transplant recipients (5.9% vs 29.9%, p=0.04). Patients with a history of malignancy were more like to die at 1 year (40.9% vs 20.6%, p=0.05), as well as patients with a history of uncontrolled diabetes (54.5% vs 19.8%, p=0.01), and patients with altered mental status (47.8% vs 17.8%, p=0.004) or respiratory symptoms (34.8% vs 17%, p=0.04). At 1-year, non-survivors had lower hemoglobin levels (10.8 ±2.4 vs 12.1±2.6 mg/dL, p=0.03), and higher HbA1c levels (7.8 ± 1.9% vs 6.1 ± 1.1 %, p=0.003). Cryptococcosis patients who died at 10-weeks and 1-year had more commonly a history of speech difficulties and ICU stay. Among cryptococcosis cases, the 10-weeks and one-year mortality was higher among those with uncontrolled diabetes: 45.5% vs 13.6%, p=0.009, and 54.5% vs 19.8%, p=0.01 respectively.

**Table 2.** Patient clinical characteristics among survivors and non-survivors at 10-weeks.

| Patient characteristics<br>(%, mean ± SD) | N | Cases survivors<br>at 10 weeks<br>(n=78) | Cases non-survivors<br>at 10 weeks (n=18) | P-values** |
| --- | --- | --- | --- | --- |
| **Type of infection** | 96 |  |  |  |
| Cryptococcal meningitis |  | 36 (46.2%) | 11 (61.1%) | 0.818 |
| Pulmonary disease |  | 30 (38.5%) | 5 (27.8%) |  |
| Skin and others |  | 7 (9%) | 1 (5.6%) |  |
| Asymptomatic antigenemia |  | 5 (6.4%) | 1 (5.6%) |  |
| **Symptoms at presentation** |  |  |  |  |
| Headaches | 219 | 27 (34.6%) | 5 (29.4%) | 0.681 |
| Altered mental status | 221 | 14 (18%) | 9 (50%) | 0.004 |
| Respiratory | 220 | 32 (41%) | 11 (61.1%) | 0.122 |
| **Labs** |  |  |  |  |
| Hemoglobin (g/dL) | 215 | 12.1 ± 2.6 | 10.4 ± 2.0 | 0.007 |
| Platelets ($10^9$/L) | 214 | 216 ± 92.1 | 151 ± 161 | 0.025 |
| Creatinine (mg/dl) | 208 | 1.3 ± 0.8 | 1.5 ± 1.1 | 0.309 |
| CD4 count (cells/μL) | 86 | 155 ± 250.8 | 202 ± 409 | 0.679 |
| HbA1c (%) | 96 | 6.2 ± 1.1 | 7.8 ± 2.0 | 0.005 |
| **CSF profile** |  |  |  |  |
| Opening pressure (cm H<sub>2</sub> O) | 54 | 28.4 ± 11.6 | 14.8 ± 1.9 | 0.055 |
| CSF WBC ($10^6$/L) | 131 | 447 ± 2376 | 55.4 ± 83.1 | 0.589 |
| CSF glucose (mg/dL) | 130 | 43.9 ± 21 | 73 ± 58.3 | 0.009 |
| CSF protein (mg/dL) | 132 | 121 ± 193 | 120 ± 159 | 0.988 |
| **Outcome** |  |  |  |  |
| ICU stay | 53 | 9 (20%) | 8 (100%) | <0.001 |
| Overall Death | 221 | 19 (24.4%) | 18 (100%) | <0.001 |
| Death within 10 weeks | 221 | 0 (0%) | 18 (100%) | NA |
| Death within 1 year | 221 | 6 (7.7%) | 18 (100%) | <0.001 |
| VP shunt | 83 | 3 (4.0%) | 1 (11%) | 0.351 |
| Cognitive deficits | 78 | 12 (16.4%) | 2 (40%) | 0.184 |
| Hearing impairment | 78 | 6 (8.1%) | 0 (0%) | 0.553 |
| Speech difficulties | 80 | 2 (2.7%) | 1 (20%) | 0.048 |
| Muscle weakness | 79 | 18 (24%) | 1 (25%) | 0.964 |
| Stroke | 34 | 5 (19.2%) | 3 (37.5%) | 0.287 |

Unadjusted Cox proportional hazard model found an increased rate of death for uncontrolled diabetes at 10-weeks (hazard ratio 3.7, CI: 1.3-10.8, p=0.01), and 1-year (hazard ratio 3.4, CI: 1.3-8.8, p=0.01) (figure 1). After adjustment for gender, age, and case/control; for every 1% increase in A1c levels, the odds of mortality increased by 40% (OR = 1.4, CI 95%: 1.0-1.9, p= 0.045). The Scatter plot showed some degree of linearity and the ANOVA R-squared/Adjusted R-squared was 95% and 6% respectively.

**Figure 1.**
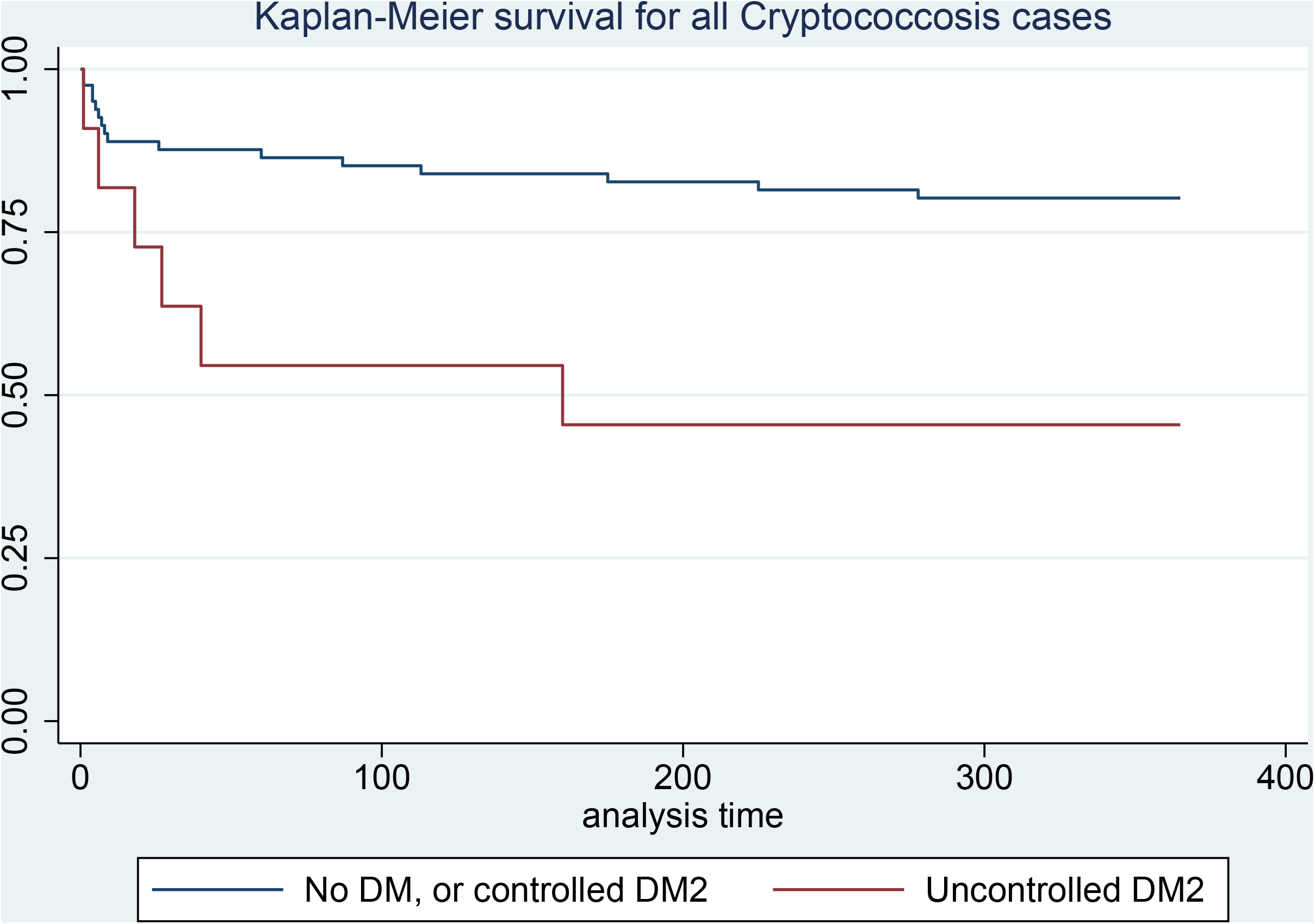
Survival curves at 1-year of Cryptococcosis cases by controlled or uncontrolled Diabetes Mellitus.

The multivariable analysis adjusted by gender, age, case/control, common risk factors for Cryptococcosis (solid organ transplantation, steroid use, HIV infection, and smoking history), and factors associated with increased mortality (altered mental status and positive blood cultures for *Cryptococcus*), revealed an independent risk of death with uncontrolled diabetes (HbA1c>7%) at 10-weeks (OR=3.6, CI: 1.1-12.3, p=0.037) and 1-year (OR=6.6, CI: 2.0-21.4, p=0.002) (Table 3).

**Table 3.** Multivariable analysis of mortality predictors.

**10-weeks**
| Variable | Odds ratio | Confidence Intervals | P-value |
| --- | --- | --- | --- |
| Uncontrolled DM* | 3.6 | 1.1-12.3 | 0.03 |
| Malignancy | 3.4 | 13.-9.1 | 0.01 |
| Cryptococcosis case | 4.0 | 1.5-10.5 | 0.005 |
| Altered Mental Status | 1.4 | 0.8-5.2 | 0.157 |

**1-year**
| Variable | Odds ratio | Confidence Intervals | P-value |
| --- | --- | --- | --- |
| Malignancy | 6.8 | 2.8-16.4 | 0.0001 |
| Uncontrolled DM* | 6.6 | 2.0-21.4 | 0.002 |
| Positive Blood cultures** | 2.6 | 0.8-8.5 | 0.106 |
| HIV | 1.9 | 0.8-4.5 | 0.159 |

D.M.: Diabetes Mellitus, \*HbA1c>7%, \*\*For *Cryptococcus spp.*

The propensity score matching with the exposure confounders revealed an ATE of 0.3 (p=0.03), and 0.4 (p=0.0001) at 10-weeks and 1-year respectively for uncontrolled diabetics. Expressing these results on a percentage scale, the chance of dying at 10-weeks and one-year is higher by 30% and 40% percentage points for uncontrolled diabetics. A multivariable sensitivity analysis of the cohort —excluding controls with diabetes but without substantial hyperglycemia (HbA1c < 7%)— showed also an independent risk of death with uncontrolled diabetes (HbA1c>7%) at 10-weeks (OR=4.3, CI: 1.3-14.4, p=0.02) and 1-year (OR=6.8, CI: 2.1-22.9, p=0.002). The MV analysis also showed a significant increase in 10-weeks (OR: 6.0, CI: 1.5-24.1, p=0.012) and 1-year mortality (OR: 4.4, CI: 1.5-13.0), p=0.008) using a different HbA1c cutoff (HbA1c >8%) as uncontrolled diabetes.

### Clinical factors associated with an increase of 10-weeks and 1-year mortality from Pulmonary Cryptococcosis and meningitis

For cryptococcal meningitis cases only, the mortality was higher for those with uncontrolled diabetes at 10 weeks (50% vs 19.1%, p=0.289) and one-year (50% vs 26.2%, p=0.46), although not statistically significant. Cases with pulmonary cryptococcosis had a higher mortality at 10 weeks (50% vs 7%, p=0.006) and one year (66.7% vs 13.8%, p=0.005)

Unadjusted Cox proportional hazard model found an increased rate of death for uncontrolled diabetes at 10-weeks (hazard ratio 8.4, CI: 1.4-50.8, p=0.02), and 1-year (hazard ratio 7.0, CI: 1.7-28.4, p=0.007) among pulmonary cryptococcosis cases (figure 2).

**Figure 2.**
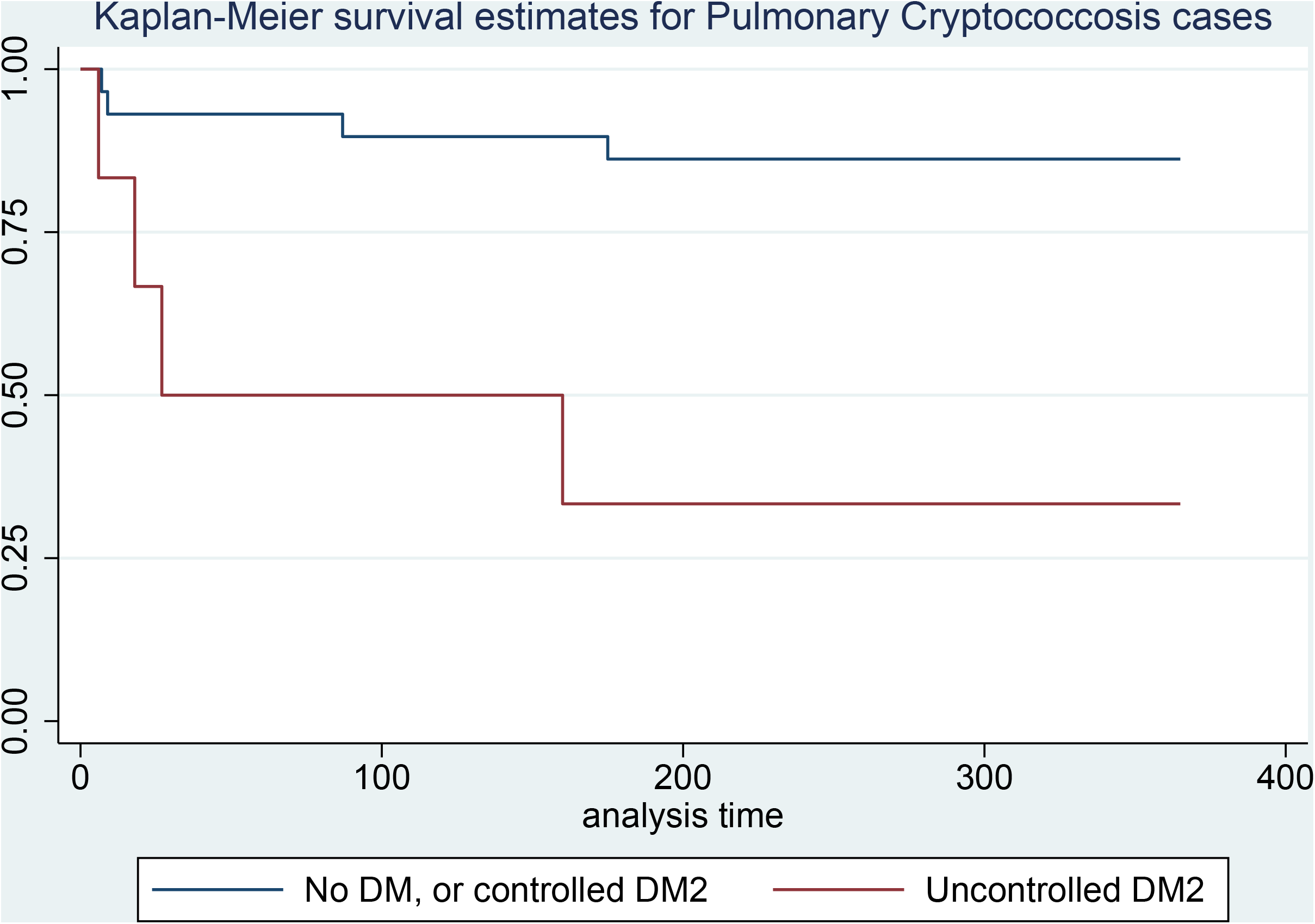
Survival curves at 1-year of Pulmonary Cryptococcosis cases by controlled or uncontrolled Diabetes Mellitus.

Multivariable analysis showed a significantly increased odds of 10-weeks (OR=4.3, CI: 1.1-16.5, p=0.035) and one-year (OR=5.9, CI: 2.2-15.8, p=0.014) mortality for uncontrolled diabetes among pulmonary cryptococcosis cases only. Malignancy was also independently associated with increased mortality.

### Clinical features and outcomes of cryptococcosis cases with diabetes and uncontrolled diabetes

Diabetes was the only known risk factor in 6 cases (6.3%), with 4 of these cases complicated by chronic kidney disease; diabetes coexisted with additional risk factors in 19 cases (19.8%). Mean HbA1c values were 5.4%, 6.9%, and 8.5% among cases without diabetes, cases with diabetes and additional risk factors, and cases with diabetes only as a risk factor, respectively. Cases with diabetes as a risk factor were older and had higher BMIs. They also were more likely to have a solid organ transplant as a risk factor, and less likely to be HIV-positive or current smokers. HbA1c levels were higher than 7% in 31.4% of the cryptococcosis cases. Labs showed a statistically significantly lower mean of hemoglobin levels (p=0.007) and an increase in the mean of creatinine levels (p=0.002) among cryptococcal cases with diabetes. Cases with diabetes also had a non-statistically higher rate of paucicellular CSF values. Only one of the six cases with only diabetes as a risk factor had disseminated disease or cryptococcal meningitis. Patients with diabetes showed no statistically significant higher rates of ICU stay, death within 10-weeks, or death within one year, but they have an increased overall death rate (any death recorded during the study period) (60% vs 31%, p= 0.01, rate ratio 2.5, CI: 1.3-4.8, p=0.007). Stroke as a complication was seen more often in patients with diabetes, but this association was not statistically significant.

Uncontrolled diabetes patients had a lower rate of HIV infection (9% vs 45%, p=0.02), were older (64 ± 8.8 vs 52 ± 14.6, p=0.008), and had most likely a pulmonary cryptococcal infection (54% vs 38.8%, p=0.02). Their BMI was also higher than controlled or non-diabetics, although this was not statistically significant.

## Discussion

We found in this single retrospective case-control study of 35 patients with pulmonary Cryptococcosis an association of uncontrolled diabetes (HbA1c >7%) with an increase of 10-weeks and 1-year mortality. These findings remained after adjusting for commonly known Cryptococcosis risk factors and other factors associated with disease mortality. Although the association remained for all Cryptococcocosis cases, the effect seems to be driven mainly by the pulmonary cases.

Survival curves demonstrated a decreased chance of survival among patients with Cryptococcosis—and also pulmonary cryptococcosis— and uncontrolled diabetes compared to Cryptococcosis patients without diabetes or with controlled diabetes. This difference was evident early in the diagnosis. Death from Cryptococcosis among people with diabetes in our cohort approached 20% at 10-weeks and 22% at 1-year, which was comparable with other cohorts. Uncontrolled diabetes correlates with increased age, end-organ failure, the presence of comorbidities, and a suboptimal immune response. Poor glycemic control associates with increased rates of serious infections and infection-related hospitalizations (19). These factors could play a role in the observed increased mortality in Cryptococcosis.

We also found diabetes to be common comorbidity among patients with *Cryptococcus* infection, and it often accompanied other established risk factors, particularly transplant status and malignancy. Diabetes as the only identifiable risk factor was relatively uncommon, although it was frequently complicated by chronic kidney disease. A recent case series identified cryptococcal meningitis as a complication of patients with nephrotic syndrome (20). The presence of diabetes mellitus translated into an observed increased chance of overall death, but it was not associated with an increase in 10-weeks or 1-year mortality.

Other U.S. cryptococcosis cohorts have reported a prevalence of diabetes of around 20% (21), similar to ours of 25%. These numbers correlate with the number of people living with diabetes based on age in the U.S. (1). An Argentinian study found a marked increase in mortality in Cryptococcal meningitis patients with AIDS and diabetes compared to those with AIDS without diabetes (85.7% vs 21.4%) (22). Matched cohort studies have identified a 20% increase in the risk of any infection among patients with diabetes (23). This risk is modulated by increasing HbA1c levels (19). Since compromised cell-mediated immunity is present in most patients affected with Cryptococcosis, one possible mechanistic explanation is the altered function of CD8^+^ T-cells and natural killer cells in patients with diabetes (24). Decreased macrophage and cytokine function (such as interleukin-12) may play a role as well (25, 26). Patients with diabetic nephropathy may also confer additional immune response impairments. Reports have shown impairments in cell-mediated immunity in chronic kidney disease (27), including decreased CD4 and CD8 cell lines (28) and T-cell proliferation (29). Solid organ transplant recipients often receive steroids as part of their immunosuppressive regimens, which in turn increases the risk to develop diabetes. The presence of diabetes in invasive infections often increases the risk of multiorgan injury and death (30).

Some cohorts haven’t found diabetes as a predictor of mortality with Cryptococcosis (6, 31), but those cohorts didn’t list the HbA1c levels, which is more specific for diabetes-associated complications. However, in a case series of 30 cases of Cryptococcosis in Diabetics in mainland China, 57% of patients didn’t list an additional underlying risk factor, and 40% had non-disseminated pulmonary cryptococcosis (32). A case-control study in Taiwan found HIV-negative patients with Cryptococcosis were more likely to have diabetes (OR: 1.5), and the presence of diabetes was also associated with an increase in 1-year mortality from Cryptococcocosis (33). Finally, a case series in Japan found diabetes mellitus in 32% of patients with pulmonary cryptococcosis (17).

We also found that non-survivors at 1-year had lower hemoglobin levels and higher HbA1c levels. The relationship between HbA1c levels and mortality may not be completely linear. There is evidence to suggest that HbA1C values are affected by changes in the erythrocyte lifespan. Iron-deficiency anemia, which increases the lifespan of erythrocytes resulting in increased glycation, is associated with falsely elevated HbA1C values. Other types of anemia, like hemolytic anemia, decrease the lifespan of erythrocytes. This is associated with falsely decreased HbA1C levels (34, 35). In this study, uncontrolled diabetes (HbA1C level >7%) was found to be a significant predictor of mortality in patients with *Cryptococcus* infection. Since non-survivors had a mean hemoglobin level of 10.4, anemia could be a possible confounder.

Pulmonary Cryptococcosis was independently associated with an increase in 10-weeks and one-year mortality risk. Although cryptococcal meningitis had higher mortality rates at the same timepoints, it did not reach statistical difference. Possible explanations are the low number of cases studied or intrinsic factors of hosts with pulmonary cryptococcosis driving the mortality differences. Malignancy can be more common among patients with pulmonary Cryptococcosis as opposed to Cryptococcal meningitis. However, the association of mortality with uncontrolled diabetes remained after adjusting for this variable.

Another finding was that transplant recipients had lower mortality at 1-year compared to non-transplant patients. This is consistent with the literature, in which transplant status is associated with lower mortality compared to non-transplant status (36, 37). Delay in diagnosis in HIV-negative, non-transplant patients compared to transplant recipients associates with worse outcomes.

There are a few limitations to this study. The retrospective selection of data limits the reliability of the predictors and limits the number of variables analyzed. We could not control for all variables associated with Cryptococcal mortality due to missing available data in our cohort. Some selection bias may exist as different observers worked on data collection. Also, the number of cases with uncontrolled diabetes was relatively low, contributing to a lack of power. We employed measures to reduce biases by adjusting for confounders. Controls were identified by a negative cryptococcal antigen, which may not be representative of the overall at-risk population and biases towards the null underestimating of the possible effects. Finally, since data was collected over two decades and some out-of-the-state deaths might haven’t been recorded, death rates could’ve been underestimated. However, our paper has been the best exercise you can do to try to answer the role of uncontrolled diabetes mellitus in Cryptococcal mortality with limited available data. A low prevalence of these patients hampers investigations designed to establish the role of glucose levels in mortality in patients with Cryptococcosis. A definitive understanding of glucose levels with mortality in these patients necessitates large prospective studies. Collecting sufficient patients for this kind of definitive study is not feasible since it would require excessively prolonged periods of subject accrual and multi-institution enrollment. Therefore, single-institution retrospective studies can generate new hypotheses and unveil or propose new associations.

Diabetes alone is an uncommon but possible risk factor by itself for acquiring *Cryptococcus* infection. It commonly accompanies additional risk factors, and it can be complicated by chronic kidney disease. Uncontrolled diabetes in Cryptococcosis—but especially pulmonary infection— may worsen outcomes from infection, leading to increased mortality. It remains to be determined whether glucose control interventions can improve clinical outcomes in patients with cryptococcal infection, and we need follow-up studies for validation of the findings in additional external cohorts.

## Data Availability

The datasets generated and analyzed in the current study are available from the corresponding author on reasonable request.

## Funding

This research received no specific grant from any funding agency in the public, commercial, or not-for-profit sectors.

## Declaration of Conflicting Interests

The authors declare that there are no conflicts of interest.

## Authors contributions

Conceptualization and design: SA, SS, AHM; data acquisition: SA, PC, KS, AHM; data analysis and interpretation: AHM, CFP, IS; original draft writing: SA, AHM; original draft review and editing: AG, PC, SC, WM, MB, JO, EM, DC, KD, LS, IS, CFP; statistical analysis: SS, AHM; supervision: CFP, AHM

## Notes

### Competing Interest Statement

The authors have declared no competing interest.

